# Decline in global transmission rates of COVID-19 through May 6 2020

**DOI:** 10.1101/2020.04.18.20070771

**Authors:** Ethan Romero-Severson, Nick Hengartner, Grant David Meadors, Ruian Ke

## Abstract

We analyzed COVID-19 data through May 6th, 2020 using a partially observed Markov process. Our method uses a hybrid deterministic and stochastic formalism that allows for time variable transmission rates and detection probabilities. The model was fit using iterated particle filtering to case count and death count time series from 55 countries. We found evidence for a shrinking epidemic in 30 of the 55 examined countries. Of those 30 countries, 27 have significant evidence for subcritical transmission rates, although the decline in new cases is relatively slow compared to the initial growth rates. Generally, the transmission rates in Europe were lower than in the Americas and Asia. This suggests that global scale social distancing efforts to slow the spread of COVID-19 are effective although they need to be strengthened in many regions and maintained in others to avoid further resurgence of COVID-19. The slow decline also suggests alternative strategies to control the virus are needed before social distancing efforts are partially relaxed.

## 1 Introduction

Since its initial outbreak in Wuhan, China in late 2019 and early 2020 (1), COVID-19 caused repeated rapid outbreaks across the global from February through April 2020. The extremely rapid spread of COVID-19 in China (2) does not appear to be an anomaly: the disease has shown a short doubling time (2.4-3.6 days) outside of China as well (3). As of May 21, 2020, the virus caused 5,034,458 reported infections, and 328,730 deaths globally (4). In response, most affected countries/regions have implemented strong social distancing efforts, such as school closures, working-from-home, shelter-in-place orders. As a result, the spread of COVID-19 slowed down substantially in some countries (5), leading to a flattening of the epidemic curve. As social distancing induces high costs to both society and individuals, plans to relax social distancing are discussed. However, changes in both the transmission rates and detection probabilities over time coupled with stochasticity due to reporting delays makes differentiating between truly subcritical dynamics and simply reduced transmission difficult.

In this report, we developed a deterministic-stochastic hybrid model and fitted the model to case incidence and death incidence time series data from 55 countries. Following the approach suggested by King et al. (6), we use a (partially) stochastic model and base our estimates on incidence rather than cumulative incidence data . Using both case count and death count of data allowed us to disentangle changes in surveillance intensity from changes in transmission (3). We found evidence for large decreases in the country-level transmission rates, in several of the worst-affected countries. Importantly, using data up to May 6, 2020, we computed 99% confidence intervals to test whether or not the data were consistent with subcritical dynamics (i.e. the reproductive number *R* was below 1 on May 6, 2020). Most countries showed large decreases in transmission rates over time, and more than half of studied countries have transmission rates below the epidemic threshold. On the other hand, many countries still appear to be showing rapid exponential growth. Given its highly contagious nature, COVID-19 can spread rapidly when strong social distancing measures are lifted, even partially (3). Alternative strategies that can effective control the virus are needed when social distancing measures are relaxed.

## 2 Materials and Methods

### 2.1 Data

Case count and death data were downloaded from the Johns Hopkins GitHub repository (https://github.com/CSSEGISandData/COVID-19) through May 6, 2020. Any country in the data that had more than 2000 cumulative cases and 100 deaths by May 6, 2020 was modeled. To minimize the effect of repatriated cases we started each time series on the first day when the cumulative number of cases exceeded 100. All data processing and model fitting were otherwise done on the incidence scale. To address obvious bulk-reporting issues in the data (e.g. sudden zeros in the data followed by very large numbers), we smoothed the data using Tukey’s 3-median method. Because several countries had days with a single death surrounded by days with no deaths, which the smoothing method would set to zero, days with a single death were not replaced with smoothed values. The original data and the smoothed data used for estimation are shown in supplemental figure s1.

### 2.2 Model

We model the spread of COVID-19 as a partially observed Markov process with real-valued states *S* (susceptible), *E* (exposed), *I* (infected), and R (recovered) to describe the latent population dynamics, and integer-valued states *C*_0_ (to be counted), *Y*_2_ (counted cases), *D*_0:3_ (dying), and *Y*_2_ (counted deaths) to model sampling into the data. The model and all of its parameters have time units of days. The latent population model is governed by the following ordinary differential equations,

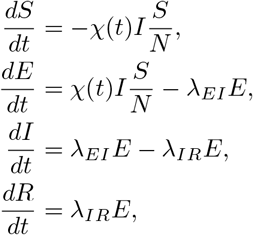

where *χ(t)* is the time-variable transmission rate and *N = S + E + I + R* is assumed to be fixed over the run of the model. λ*_EI_* is the rate at which exposed persons become infectious and *λ_IR_* is the rate at which cases recover (i.e. are either no longer infectious or die). At every time interval, we sample persons moving from the *E* to *I* states into a stochastic arm of the model that is used to calculate the likelihood of the data.

To relate the latent population model to data, we randomly sample individuals from the unobserved population into a stochastic process that models the random movement from infection to being either counted as an observed case or counted as an observed death. The number of persons sampled into the observation arm of the model over a time interval *dt* is given by *X = (X*_1_, *X*_2_, *X*_3_, *X*_4_) ~ Multinomial(*g(Eλ_EI_dt*), 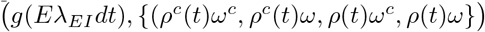 where *ρ(t)* and *ω* are the probabilities that an infected person will be counted or die respectively, *ρ^c^(t)* and *ω^c^* are the probabilities of being not sampled or not dying respectively, and *g(u)* is a stochastic function that maps from real-value *u* to integer *g*⌊*u*⌋; it takes value ⌈*u*⌉ with probability *u* mod 1 and value ⌈*u*⌉ with probability 1 − (*u* mod 1). In plain English, the model tracks the random fate of each newly infected person as they move from the exposed to infected state with respect to eventually being observed as a case and/or a death in data. At each time step of length *dt*, the change in state space is given by,

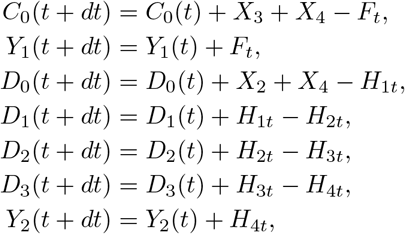

where *F_t_*|*C*_0_ is a random variate from Bin(*C*_0_, 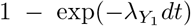), and *H_it_* is a random variate from Bin (*D*_i−1_, 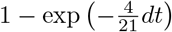). *X_i_* indicates the *i^th^* element of Multinomial random variate defined above. The rate λ*_Y_*_1_ determines the rate at which persons who will be counted are counted (i.e. lower values of λ*_Y_*_1_ mean a longer delay in cases showing up in the data). The values of *Y*_1:2_ are set to zero at the beginning of every day such that they accumulate the simulated number of cases and deaths that occur each day. Both the transmission rate and detection probability are allowed to vary with time in the following way:

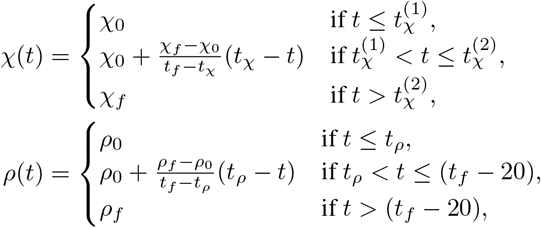

where *t_f_* is the final time in the given dataset. In plain English, the transmission rate is constant up to some time, 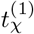 where it linearly increases or decreases to the value *χ_f_* by time 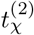. The model is constrained such that 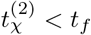, that is, the transmission rate must be constant for at least 20 days before the end of the data collection period. This constraint is in place to avoid overfitting the final transmission rate. The detection rate likewise has a linear increase or decrease beginning at time *t_ρ_;* however, the increase or decrease continues to a fixed point that is 20 days before the final datum. Variation in the testing rate over the course of the epidemic can strongly bias estimates of the population growth rate (derivation for the exponential growth case in supplemental Material); not allowing the detection probability to change over time could lead to discordance in the case count and death count time series.

### 2.3 Parameter estimation

We assume that the data are Negative Binomial-distributed, conditional on the simulated number of cases and deaths that occur in a given time interval, i.e. the number of cases in the *i^th^* observation period has density NegBin(*Y*_1_*, ϵ*_1_) and the number of deaths in the *i^th^* observation period has density NegBin(*Y*_2_, *ϵ*_2_). The Negative Binomial is parameterized such that the first argument is the expectation and the second is an inverse overdispersion parameter that controls the variance of the data about the expectation; as *ϵ_i_* becomes large, the data model approaches a Poisson with parameter *Y_i_*. Both *ϵ*_1_ and *ϵ*_2_ were estimated from the data.

Parameter estimation had two distinct steps: model selection and computation of confidence intervals. In the model selection phase, the model was fit to the data using an iterated particle-filtering method, implemented in the `pomp` R library (7). To optimize the likelihood of the data, we used 1500 particles in 125 iterations for each country. The reported likelihoods were measured using 4500 particles at the optimized Maximum Likelihood Estimates (MLEs). The Monte Carlo error in the likelihood measurement was less than 0.1 log units.

For all fits, the initial state at time zero is computed by assuming there were *I*(0) infected persons, 21 days before the first reported death (by definition time one). The number of initial number of susceptibilities was assumed to be the predicted 2020 population size of the given country (8). The model is then simulated forward for 21 days, assuming exponential growth with transmission rate χ_0_, which is taken at the initial state of the model at time zero. Parameters have constraints *I*(0) ∈ [1,1000], *ρ*_0_ ∈ [0,1], *ρ_f_* ∈ [0,1], 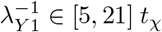 and *t_ρ_* ∈ [0*,t_f_* − 10]; all other parameters are constrained to be positive.

Because the data were highly variable in the complexity of the patterns they showed, we considered three nested models of increasing complexity for each country. The first model (model 1) assumed simple exponential growth with a constant sampling probability, i.e. *ρ*_0_ = *ρ_f_* and 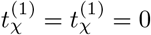. This amounts to a period of exponential growth withtransmission rate *χ*_0_ in the pre-data period and then a constant transmission rate *χ_f_* over the observation period. The second model (model 2) allowed the transmission rate to vary but kept the detection probability constant, i.e. *ρ*_0_ = *ρ_f_*. The third model (model 3) allowed both the transmission rate and detection rate to vary, i.e. all parameters estimated. We determined the best model for each country by a sequence of likelihood ratio tests first comparing model 1 and model 2 and then model 2 to model 3.

Because we are using an optimization method, we do not have direct access to samples from the likelihood surface directly. To obtain estimates of the parametric uncertainly in the final transmission rate we computed 99% confidence intervals using the profile likelihood method (9). We used the 99% confidence level to minimize the effect of numerical noise coming from both the optimization procedure and the Monte Carlo error in the measurement of the likelihood function.

## 3 Results

We fit our model to data collected from 55 countries. Model fits are shown in Figure 1, and parameter values are given in Table 5. The model can capture the data well, with a few exceptions. The model was not able to find a robust fit to the data from Bangladesh; in general, the upside down ‘V’ shape to the deaths could not be captured well in such a short time series. Algeria had a similarly odd pattern in the deaths time series that the model could not capture in detail, although the overall trend in deaths was recovered. In previous versions of this paper, the model had a hard time fitting data from both Italy and Spain. However, given the longer time series and modifications to the model form, we now find that both Italy and spain are well-captured by the model.

**Figure 1:**
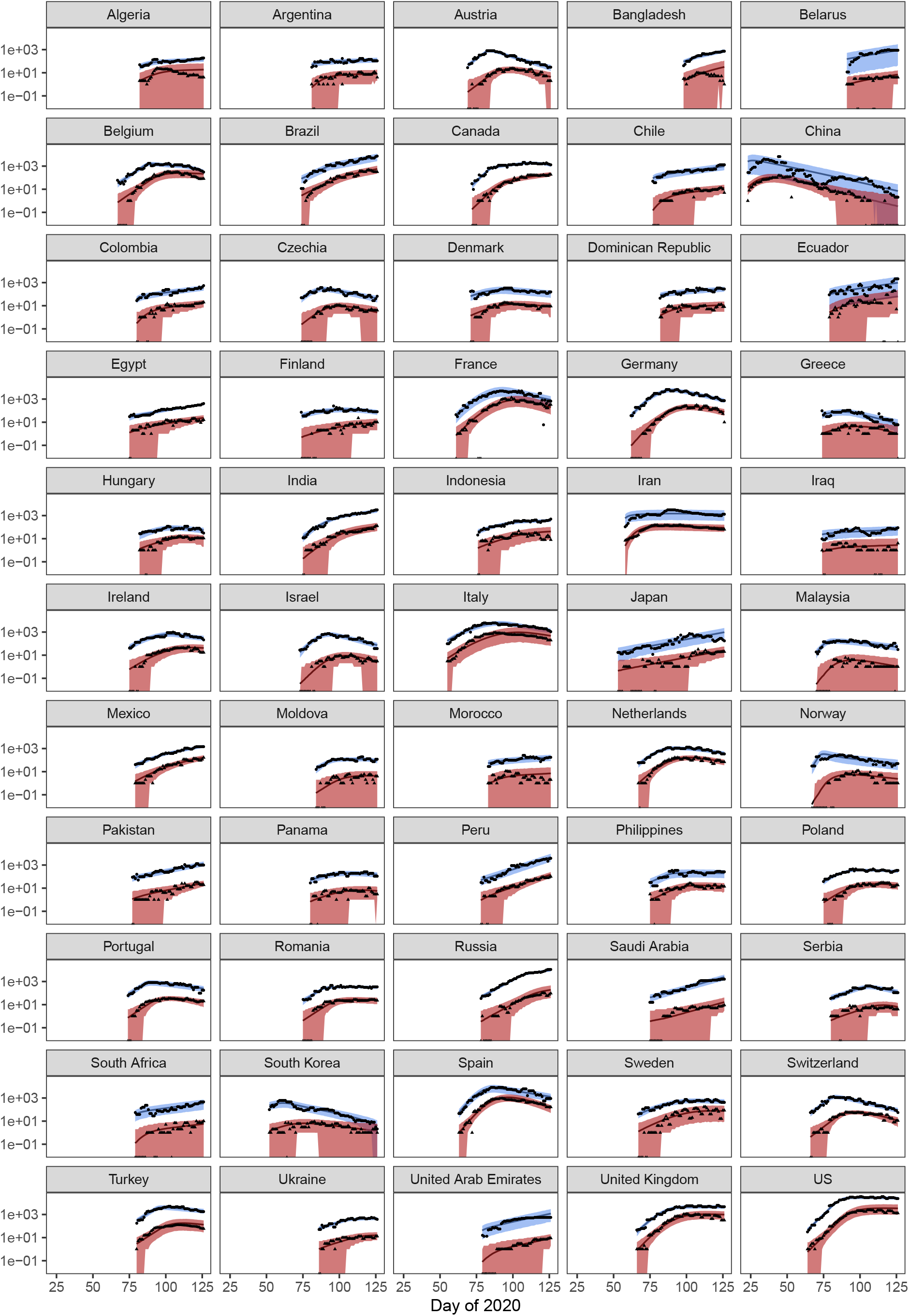
Model fits. Smoothed data points are shown in black and the expectations from the model are shown as solid lines. The envelope shows the 95% CIs of the distribution of the data conditional on the maximum likelihood model fit for the best model for each country. The x-axis is measured in sequential days of the year 2020.

**Table 1:** Table of key parameter values. The ‘Model’ column referrers to the best model defined in the model selection procedure. The values *r*_0_ and *r_f_* are the initial and final exponential growth rates implied by the other model parameters. The growth rate *r* and transmission rate *χ* are derived according to (11). *χ*_0_, *χ_f_*, *r*_0_, *r_f_*, have day^−1^ units; *ρ*_0_ and *ρ_f_* are probabilities.

| Country | Model | $\chi_f$ | CI $\chi_f$ | $r_f$ | CI $r_f$ | $\rho_0$ | $\rho_f$ |
| --- | --- | --- | --- | --- | --- | --- | --- |
| Algeria | 2 | 0.111 | 0.109, 0.117 | 0.008 | 0.007, 0.013 | 0.08 | 0.08 |
| Argentina | 1 | 0.111 | 0.11, 0.123 | 0.008 | 0.008, 0.017 | 0.16 | 0.16 |
| Austria | 2 | 0.024 | 0.022, 0.025 | -0.07 | -0.072, -0.069 | 0.25 | 0.25 |
| Bangladesh | 3 | 0.197 | 0.195, 0.199 | 0.065 | 0.064, 0.066 | 0.1 | 0.1 |
| Belarus | 1 | 0.163 | 0.119, 0.223 | 0.044 | 0.014, 0.08 | 0.79 | 0.79 |
| Belgium | 3 | 0.092 | 0.088, 0.095 | -0.006 | -0.009, -0.004 | 0.05 | <0.01 |
| Brazil | 2 | 0.185 | 0.183, 0.186 | 0.058 | 0.057, 0.059 | 0.05 | 0.05 |
| Canada | 3 | 0.156 | 0.154, 0.157 | 0.039 | 0.038, 0.04 | 0.11 | <0.01 |
| Chile | 1 | 0.154 | 0.153, 0.156 | 0.038 | 0.037, 0.039 | 0.55 | 0.55 |
| China | 1 | 0.025 | 0.023, 0.027 | -0.069 | -0.071, -0.067 | 0.24 | 0.24 |
| Colombia | 1 | 0.15 | 0.137, 0.154 | 0.035 | 0.027, 0.038 | 0.18 | 0.18 |
| Czechia | 2 | 0.027 | 0.009, 0.039 | -0.066 | -0.088, -0.054 | 0.28 | 0.28 |
| Denmark | 2 | 0.05 | 0.044, 0.061 | -0.042 | -0.048, -0.032 | 0.19 | 0.19 |
| Dominican Republic | 1 | 0.122 | 0.12, 0.136 | 0.016 | 0.015, 0.026 | 0.24 | 0.24 |
| Ecuador | 1 | 0.163 | 0.134, 0.194 | 0.044 | 0.025, 0.063 | 0.13 | 0.13 |
| Egypt | 2 | 0.165 | 0.163, 0.165 | 0.045 | 0.044, 0.046 | 0.12 | 0.12 |
| Finland | 3 | 0.131 | 0.128, 0.133 | 0.023 | 0.021, 0.024 | 0.23 | 0.03 |
| France | 2 | 0.002 | <0.001, 0.004 | -0.097 | -0.1, -0.095 | 0.07 | 0.07 |
| Germany | 3 | 0.023 | 0.019, 0.057 | -0.071 | -0.076, -0.036 | 0.38 | 0.02 |
| Greece | 2 | 0.024 | <0.001, 0.048 | -0.07 | -0.1, -0.044 | 0.18 | 0.18 |
| Hungary | 2 | 0.087 | 0.077, 0.093 | -0.011 | -0.019, -0.005 | 0.08 | 0.08 |
| India | 3 | 0.158 | 0.156, 0.16 | 0.041 | 0.04, 0.042 | 0.19 | 0.05 |
| Indonesia | 2 | 0.122 | 0.116, 0.123 | 0.017 | 0.012, 0.017 | 0.07 | 0.07 |
| Iran | 1 | 0.092 | 0.09, 0.093 | -0.007 | -0.008, -0.005 | 0.15 | 0.15 |
| Iraq | 1 | 0.112 | 0.105, 0.118 | 0.009 | 0.004, 0.014 | 0.21 | 0.21 |
| Ireland | 2 | 0.069 | 0.063, 0.081 | -0.025 | -0.03, -0.015 | 0.12 | 0.12 |
| Israel | 2 | 0.012 | 0.002, 0.026 | -0.085 | -0.097, -0.067 | 0.66 | 0.66 |
| Italy | 2 | 0.051 | 0.049, 0.054 | -0.042 | -0.043, -0.039 | 0.06 | 0.06 |
| Japan | 1 | 0.18 | 0.177, 0.183 | 0.054 | 0.053, 0.056 | 0.18 | 0.18 |
| Malaysia | 2 | 0.043 | 0.037, 0.045 | -0.049 | -0.056, -0.047 | 0.54 | 0.54 |
| Mexico | 2 | 0.175 | 0.174, 0.192 | 0.052 | 0.051, 0.062 | 0.05 | 0.05 |
| Moldova | 3 | 0.085 | 0.079, 0.103 | -0.012 | -0.017, 0.003 | 0.3 | <0.01 |
| Morocco | 1 | 0.121 | 0.12, 0.125 | 0.016 | 0.015, 0.019 | 0.23 | 0.23 |
| Netherlands | 3 | 0.046 | 0.043, 0.048 | -0.046 | -0.049, -0.045 | 0.08 | 0.01 |
| Norway | 2 | 0.063 | 0.058, 0.07 | -0.03 | -0.036, -0.024 | 0.35 | 0.35 |
| Pakistan | 1 | 0.183 | 0.176, 0.194 | 0.056 | 0.052, 0.063 | 0.34 | 0.34 |
| Panama | 2 | 0.061 | 0.059, 0.084 | -0.032 | -0.034, -0.012 | 0.34 | 0.34 |
| Peru | 1 | 0.255 | 0.255, 0.271 | 0.097 | 0.097, 0.106 | 0.1 | 0.1 |
| Philippines | 2 | 0.082 | 0.081, 0.084 | -0.015 | -0.015, -0.013 | 0.16 | 0.16 |
| Poland | 2 | 0.071 | 0.068, 0.072 | -0.024 | -0.027, -0.023 | 0.19 | 0.19 |
| Portugal | 2 | 0.046 | 0.043, 0.049 | -0.046 | -0.05, -0.044 | 0.22 | 0.22 |
| Romania | 2 | 0.084 | 0.076, 0.085 | -0.013 | -0.019, -0.012 | 0.15 | 0.15 |
| Russia | 2 | 0.148 | 0.146, 0.15 | 0.034 | 0.033, 0.036 | 0.43 | 0.43 |
| Saudi Arabia | 1 | 0.222 | 0.218, 0.227 | 0.079 | 0.077, 0.082 | 0.96 | 0.96 |
| Serbia | 2 | 0.001 | <0.001, 0.005 | -0.099 | -0.1, -0.094 | 0.41 | 0.41 |
| South Africa | 1 | 0.148 | 0.133, 0.151 | 0.034 | 0.024, 0.036 | 0.5 | 0.5 |
| South Korea | 3 | 0.059 | 0.059, 0.061 | -0.034 | -0.035, -0.033 | 0.45 | 0.01 |
| Spain | 2 | 0.014 | 0.005, 0.027 | -0.082 | -0.093, -0.067 | 0.08 | 0.08 |
| Sweden | 3 | 0.169 | 0.093, 0.182 | 0.048 | -0.006, 0.056 | 0.08 | 0.07 |
| Switzerland | 2 | 0.024 | 0.023, 0.026 | -0.069 | -0.071, -0.067 | 0.17 | 0.17 |
| Turkey | 2 | 0.059 | 0.058, 0.061 | -0.034 | -0.035, -0.032 | 0.22 | 0.22 |
| Ukraine | 2 | 0.085 | 0.082, 0.101 | -0.012 | -0.014, 0.001 | 0.33 | 0.33 |
| United Arab Emirates | 1 | 0.193 | 0.19, 0.23 | 0.063 | 0.061, 0.084 | 0.99 | 0.99 |
| United Kingdom | 3 | 0.095 | 0.094, 0.097 | -0.004 | -0.004, -0.002 | 0.05 | <0.01 |
| US | 3 | 0.1 | 0.098, 0.101 | 0 | -0.001, 0.001 | 0.09 | 0.07 |

Overall, the model slightly overestimates the number of deaths around the time where deaths begin to decrease. However, this was generally corrected if the time series was long enough. Including temporal heterogeneity in the time from infection to detection of a COVID-19 death would likely correct this; however, it is not clear that this is advisable, as death counts are likely under-reported.

All countries except Japan and saudi Arabia were found to have lower transmission rates on May 6, 2020 than at the beginning of the observation period, suggesting a global decline in the transmission of COVID-19 though May 6, 2020. However, the initial transmission rate should be interpreted cautiously, as we allowed a wide range of infected persons to exist 21 days before the first observation. That is, the initial transmission rate parameter is rather a convolution of the unknown number of infected persons and initial growth rate consistent with the data.

We found significant evidence for subcritical dynamics in 27 countries (3 countries had subcritical point estimates but their CIs contained the epidemic threshold). Figure 2 shows the point estimates and CIs of the final transmission rate on May 6, 2020 for all countries, stratified by continent (10). European countries had the highest probability of being subcritical (21 of 25) with Asia (7 of 15) and the Americas (2 of 11) having fewer subcritical countries. None of the countries in Africa that met the inclusion criteria had subcritical dynamics.

**Figure 2:**
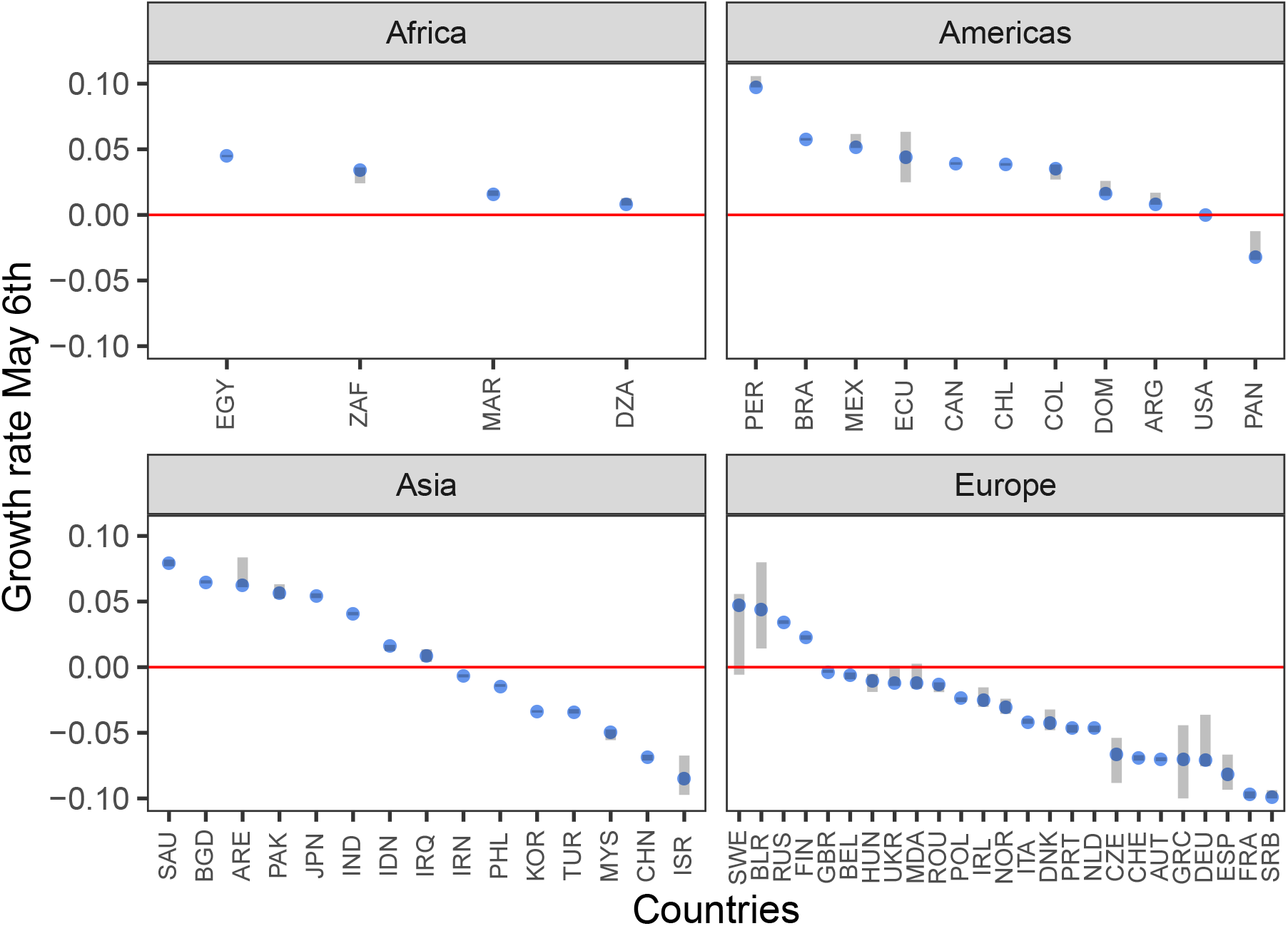
Values of the final exponential growth rates as of May 6 2020. Blue dots indicate point estimates and grey bars indicate the 99% confidence intervals. The red line delineates exponential growth from exponential decay. Countries below this line have a shrinking epidemic. Country names are indicated by ISO Alpha-3 codes.

Figure 3 shows the predicted number of deaths projected out to July 5, 2020 assuming that all parameter values are constant over the period May 6 through July 5 2020. The average duration of the country-level epidemic in European countries is longer than in Asia, leading to a higher level of death, despite Asian countries having on-average higher growth rates. However, in the Americas, the predicted deaths are higher with 8 of 11 countries having total predicted deaths greater than 0.1% of the total population by July 5, 2020. The model predicts 1,320,170 total COVID-19 deaths in all 55 countries on July 5, 2020; of those deaths, 21% are predicted to occur in the US. The deaths due to COVID-19 in Europe are lower than the average number of reported deaths in a period of the same length for all countries in the data set that also had all-cause death counts from previous years. However, in the Americas, the COVID-19 death counts are approaching the all-cause death levels in several countries, suggesting that COVID-19 deaths are approaching a doubling of all-cause deaths in those countries.

**Figure 3:**
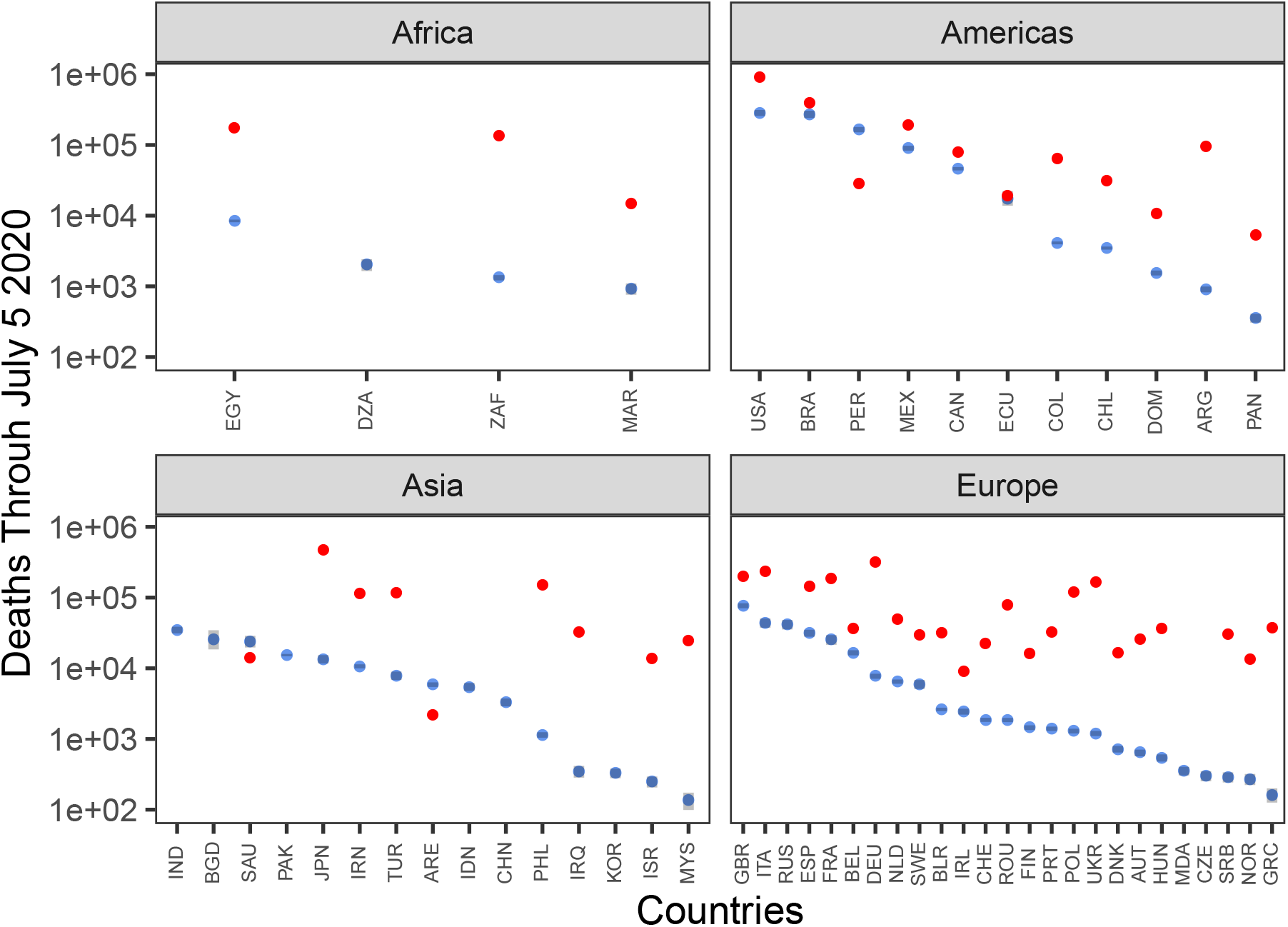
Predicted number of cumulative deaths through July 5 2020. The distribution of the total number of predicted deaths in the observation period plus 60 days assuming final model maximum likelihood values on May 6 2020 are constant though July 5 2020. Blue dots indicate median values and grey bars show the 95% confidence interval. Red dots show the average number of deaths in an average period of the same length in the World Health Organization death data from the most recent year available. The y-axis is on the log10 scale. Country names are indicated by ISO Alpha-3 codes

## 4 Discussion

Our model found evidence for reductions in transmission rates of COVID-19 in 53 of 55 examined countries. Encouragingly, of those countries, we found statistical evidence that the size of the epidemic is decreasing in 27 countries, i.e. the effective reproductive number is less than 1, using data up to May 6, 2020. This suggests that, despite the highly heterogeneous populations represented by these countries, the growth of COVID-19 outbreak can be reverted. Although our model cannot attribute exact causes to the global decline in transmissions rates, most countries implemented sustained, population-level social distancing efforts over a period of weeks to months. These efforts are highly likely to play a major role in reducing the transmission of COVID-19 (5).

We estimated that in countries with decreasing transmission, the rate of decrease is in general less than 0.1/day (average -0.04/day). Based on data from 8 European countries, the US, and China, we previously estimated that in the absence of intervention efforts, the epidemic can grow at rates between 0.19-0.29/day (2). This means that the outbreak can grow rapidly and quickly wipe gains made though public heath efforts made if social distancing measures are completely relaxed. For example, if the rate of decrease under strong public health interventions is 0.1/day and the growth rate in the absence of public heath interventions is 0.2/day, then, the number of cases averted in two weeks of intervention will be regained in only one week. Social distancing measures have their own social costs. Our results suggests alternative strategies to control the virus are needed in place when social distancing efforts are relaxed. Due to the uncertainties in the impact of each specific control measures, changes to policies should be made slowly because the signal of changing transmission can take weeks to fully propagate into current data streams as a result of the long lag between infection to case confirmation (as we estimated to be on average approximately 2 weeks).

Overall, our results suggest that COVID-19 is controllable in diverse settings using a full range of strong and comprehensive non-pharmaceutical measures, and that future deaths from the disease are avoidable.

## 5 Tables

| Symbol | Description | Value |
| --- | --- | --- |
| $\lambda_{EI}$ | Latency rate | $\frac{1}{3}$ |
| $\lambda_{IR}$ | Recovery rate | $\frac{1}{10}$ |
| $\omega$ | Case fatality ratio | 1% |
| $\chi_0$ | Initial transmission rate | fit |
| $t_\chi^{(1)}$ | Time of beginning of trans. rate change | fit |
| $t_\chi^{(2)}$ | Time of ending of trans. rate change | fit |
| $\chi_f$ | Final transmission rate | fit |
| $\rho_0$ | Initial probability of being counted | fit |
| $t_\rho$ | Time of counting prob. change | fit |
| $\rho_f$ | Final counting prob. | fit |
| $I(0)$ | Initial number infected | fit |

## 6 Supplemental

### 6.1 Estimating the infection and counting rates with exponential growth

Consider an exponential growth model early in the outbreak where infected (*I*) persons transmit at rate *β*, are counted (*C*) at rate *α(t)*, recover (*R*) at rate *γ*_1_, and die (*D*) at rate *γ*_2_. When individuals are counted, they are not removed from *I* and transmit at the same rate as uncounted persons. The differential equations for *I* and *C* cases in time *t*, where (*I, C, R, D*) are dimensionless numbers, but (*α, β, γ*_1_*,γ*_2_) have units of (time)^−1^, state,

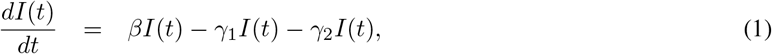

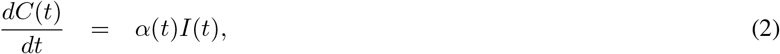

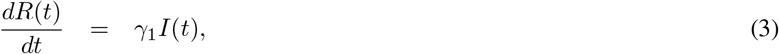

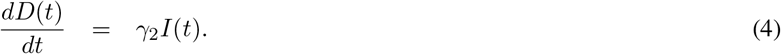

Let us also tie *C* to *R*. We have that,

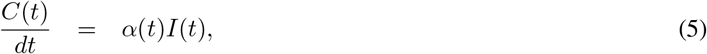

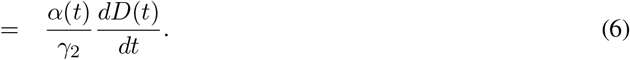

The problem is that we only observe *dC(t)/dt* and *dD(t)/dt*. So algebraically,

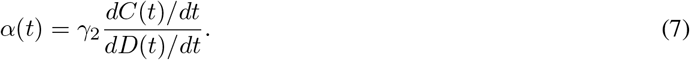

Given the model specified, it is true that *α = γ_2_(dC/dt)/(dD/dt*). Since they are total derivatives, it is also true that *α = γ*_2_*dC/dD*.

#### 6.1.1 Constant counting rate

It is a first-order problem to show that,

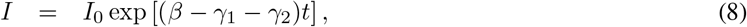

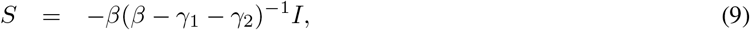

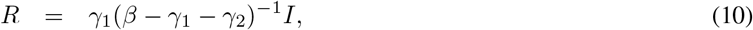

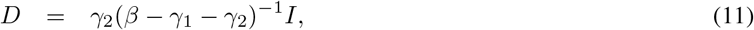

as well as, when *dα/dt =* 0,

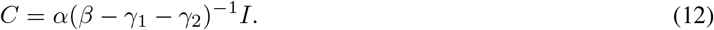

An interpretation is that *I(t) = C(t + τ)*, with time lag of *τ* applied to *t*, where,

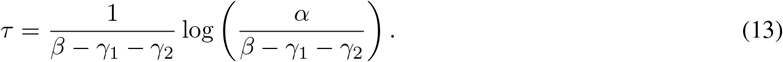

This simply shows that if the counting rate is constant, then the value of *C* is simply equal to *I* shifted in time by *τ*.

#### 6.1.2 Time-varying counting rate

For time-varying *α*, at time *t* with initial count *C*_0_ at time *t*_0_, in terms of (the only quantities observable) *C* & *D*,

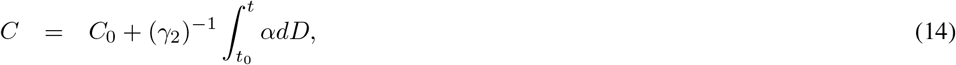

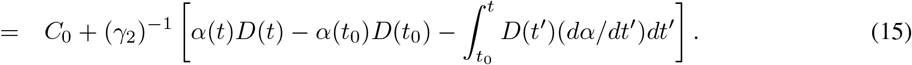

Stipulating that *C*_0_ *= (γ_2_)α(t*_0_)*D(t*_0_), up to an additive constant,

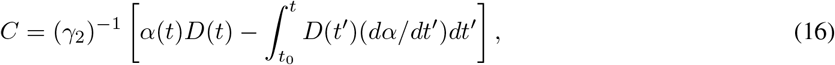

which reduces back, if *dα/dt =* 0, to the time-integral of Equation 7:

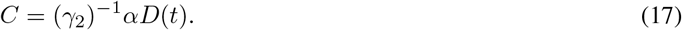

#### 6.1.3 Exponential-growth difference between counts and infections

In the case where *dα/dt >* 0, we find the difference (*d* log *C)/dt* − (*d* log *I)/dt*. It is generally true that,

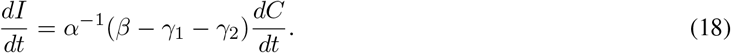

Using *dx = xd* log *x* to transform *C, I*,

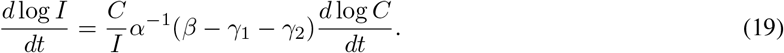

Yet integration by parts shows that, for general *dα/dt*,

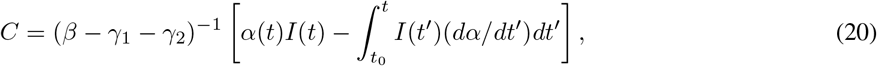

substitution of which yields,

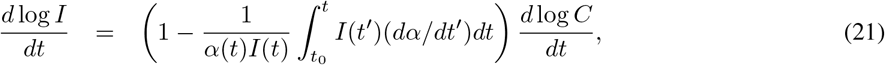

Whenever *dα/dt* is positive and, as almost always the case, *α, I, dI/dt >* 0, then Equation 21 implies that log *C* grows faster than *log I*. In the approximation that *α* changes slowly,

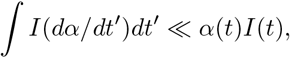

then Taylor expanding and rearranging terms yields a simpler expression,

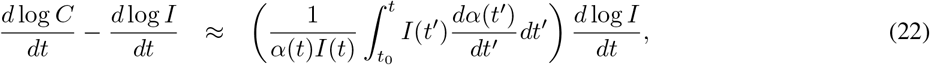

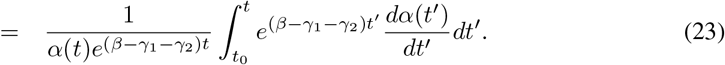

For all positive *α, dα/dt*, the right-hand side is positive. Evidently, the growth rate in logarithmic *C* exceeds that in logarithmic *I*. The growth rate of logarithmic *C* can be readily inferred from a log-linear plot, but *I* is generally unknown. When the testing rate increases during an epidemic’s exponential growth phase, the number of counts *C* increases faster than the number of infections *I*.

**Figure S1:**
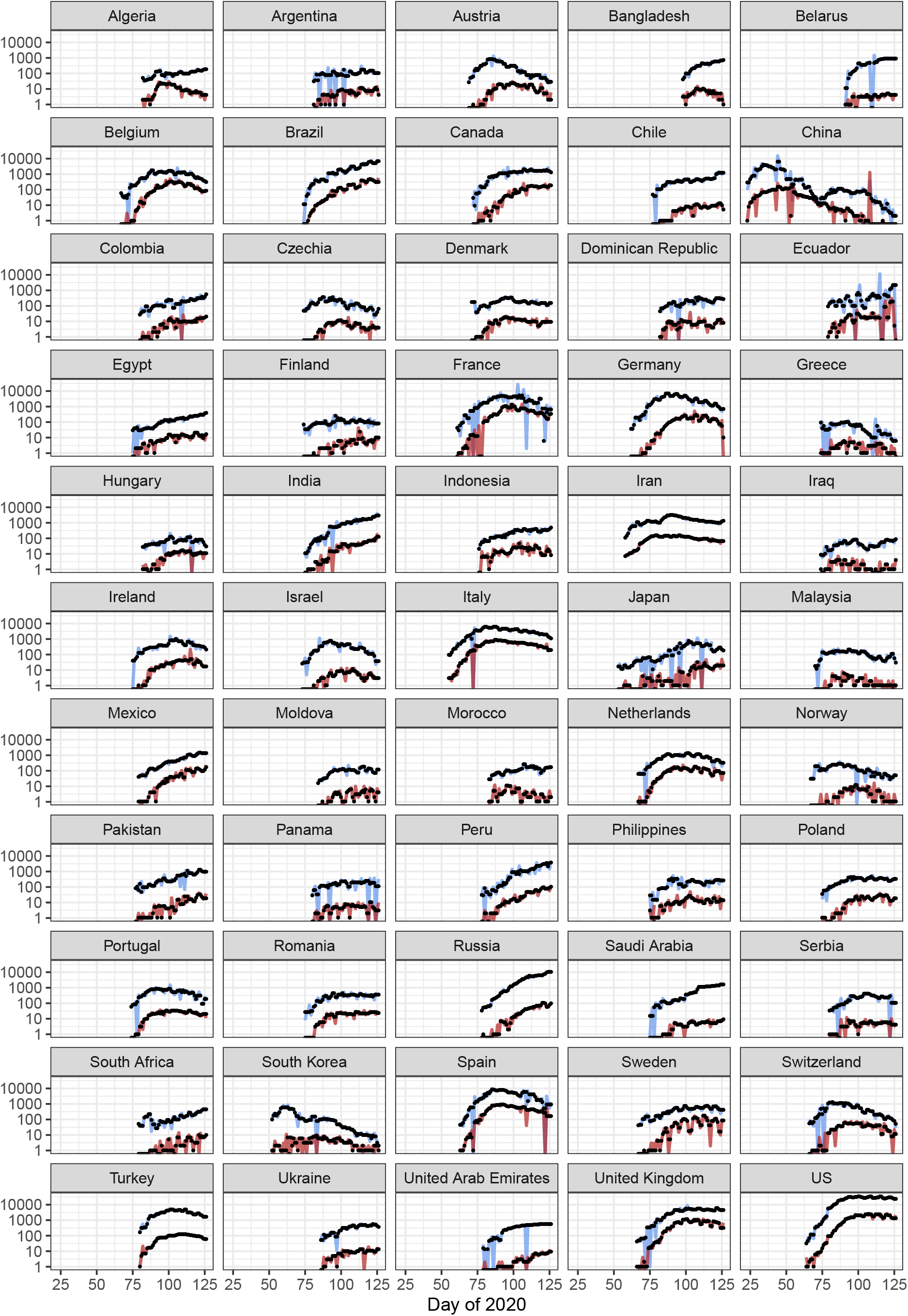
Raw and smoothed data. Raw data for cases (blue lines) and deaths (red lines) are shown with the smoothed time series (black) used for model fitting. The x-axis is measured in sequential days of 2020.

## Data Availability

All data used in the manuscript are publicly available at https://github.com/CSSEGISandData/COVID-19

